# Outcomes of patients with calcific aortic valve disease according to the extent of cardiac damage

**DOI:** 10.1101/2024.10.18.24315782

**Authors:** Matthew K Moore, Gregory T Jones, Gillian Whalley, Bernard Prendergast, Michael J.A. Williams, Sean Coffey

**Affiliations:** Department of Medicine, HeartOtago, Dunedin School of Medicine, University of Otago, Dunedin, New Zealand; Department of Surgical Sciences, Dunedin School of Medicine, University of Otago, Dunedin, New Zealand; St Thomas’ Hospital and Cleveland Clinic London, United Kingdom; Department of Cardiology, Dunedin Hospital, Te Whatu Ora, Dunedin, New Zealand

## Abstract

**Background:** A staging system for aortic stenosis (AS) based upon the extent of cardiac damage has been proposed to better stratify risk and evaluate the benefit of aortic valve intervention (AVI), especially in those with moderate AS. We sought to evaluate the prognostic value of this staging system.

**Methods:** Data from initial clinically indicated echocardiograms performed between 2010 and 2018 in patients >18 years of age were extracted and linked to national outcome data. The combined primary outcome was mortality or hospitalization with heart failure.

**Results:** Amongst 24,699 patients, 513 and 920 had moderate and mild AS, respectively. In moderate AS, Stage 0 cardiac damage was present in 9.4%, Stage 1 in 53.7%, Stage 2 in 31.1%, Stage 3 in 3.2%, and Stage 4 in 2.6%. In mild AS, rates were 11.5%, 57.8%, 25.0%, 2.6%, and 3.0% for each consecutive stage. Increasing stage was associated with increased risk of the primary outcome in both moderate (HR 1.62/stage) and mild AS (HR 1.93/stage). After censoring at the time of AVI, increasing stage was also associated with mortality in moderate (HR 1.97/stage) and mild AS (HR 2.06/stage).

**Conclusion:** Stage of cardiac damage predicts prognosis in both moderate and mild AS to a similar extent. Outcomes may therefore not be fully related to the haemodynamic consequences of valve disease, and hence may not be entirely reversible after valve intervention. Revised management algorithms focusing on earlier intervention and novel treatment strategies targeting cardiac damage are needed to improve clinical outcomes in patients with AS.

## Background

Calcific aortic valve disease (CAVD) poses an increasing challenge to health and the increasing incidence in aging populations creates a need to improve our ability to identify those who may benefit most from early intervention. Whilst current indications for intervention are largely directed by symptom status, cardiac damage is increasingly recognised in this patient cohort and associated with mortality in both moderate and severe aortic stenosis (AS) (1–4). A staging system for the classification of cardiac damage demonstrates that baseline abnormalities predict poor outcomes, whilst improved staging following aortic valve intervention (AVI) is associated with reduced mortality at two year follow up (5).

Use of this staging system to identify cardiac damage could improve outcomes following AVI compared to the current “wait for symptoms” approach and facilitate the identification of those who may benefit most from intervention. However, the utility of the system depends on its ability to clearly delineate between severe haemodynamic effects related to AS and those related to other pathologies. In this study, we aim to assess the use of this staging system in both mild and moderate AS, and clarify whether adverse outcomes are driven by the valve pathology.

## Methods

### Study cohort

The local regional health authority (Te Whatu Ora – Southern) provides secondary and tertiary cardiology services to the lower part of New Zealand’s South Island, with approximately 5000-6500 echocardiograms performed annually amongst the 330,000 inhabitants. Our cohort comprised all patients aged >18 years undergoing a clinically indicated echocardiogram at Dunedin and Invercargill Hospitals, New Zealand over a nine-year period (January 1^st^ 2010 - December 31^st^ 2018). Only those with mild or moderate AS were included in the primary analysis.

### Collection of echocardiographic data

This was a retrospective study of routinely acquired clinical information, and details of data acquisition and cleaning are fully described elsewhere (6). Data were housed in the Syngo Dynamics echocardiographic picture archiving and communication system (PACS) (version VA20F, Siemens Healthineers, Erlangen, Germany), and studies were extracted using the proprietary Syngo Dynamics Data Miner in a comma-delimited file. In all, 42,517 studies were extracted and underwent data cleaning, using previously described variable definitions and methods (6). Only the first study for each individual was included, and those with missing CAVD status (n=1,323) were excluded, leading to a final sample size of 24,699 patients who were linked by national health index (NHI) number to provide outcome data.

Qualitative categorical variables were extracted from the echocardiogram report using tailored functions to analyse free text fields for relevant variables, and the report text used to categorize CAVD status. The classification of CAVD was based on the reading cardiologist’s clinical description in the echocardiogram report. Thus, “sclerosis” was reported if the valve was described as sclerosed, thickened, or calcified, in the absence of higher levels of severity. Clinically reported mild-to-moderate and moderate-to-severe CAVD were collapsed down to mild and moderate disease, respectively. Aortic valve morphology was recorded as bicuspid if explicitly stated, and otherwise as tricuspid. Patients who had undergone previous surgical or transcatheter aortic valve intervention were described separately. Information on other cardiovascular comorbidities (such as diabetes, hypertension, or chronic kidney disease) were not available in our dataset.

### Data validation

To determine the accuracy of the Data Miner output, 100 studies were randomly selected and full data extraction was compared to the final clinical echocardiogram report in the electronic health record, which showed excellent agreement (6).

### Staging of CAVD

Stages of extra-valvular cardiac damage were classified based upon a modification of previous descriptions (1):

- **Stage 0:** no extra-valvular cardiac damage
- **Stage 1:** LV mass >224g (male) or >162g (female), E/e’ >14 or not measured, or left ventricular ejection fraction <40%
- **Stage 2:** moderate or greater left atrial dilation, atrial fibrillation, or moderate or more mitral regurgitation
- **Stage 3:** right ventricular systolic pressure >60 mmHg or moderate or more tricuspid regurgitation
- **Stage 4:** moderate or greater impairment of right ventricular systolic function

LV mass was calculated using the Deveraux formula. Body surface area was frequently not available in our dataset, so the upper limits of the normal range of absolute LV mass were used (7). Those with an LV mass >1000g were excluded since this value was unlikely to reflect a true measurement. Similarly, E/e’ was not measured in those with significant mitral valve disease, mitral annular calcification, arrhythmia or other settings where this parameter is inaccurate, and assumed to be abnormal if not recorded (8).

### Primary and secondary outcomes

The pre-specified primary outcome was a composite of mortality and hospitalisation with a primary diagnosis of heart failure (HHF). A time to first event analysis was pre-specified since multiple events were possible in an individual patient.

Secondary outcomes were:

1. Aortic valve intervention (AVI) – surgical aortic valve replacement, transcatheter aortic valve implantation or balloon aortic valvuloplasty
2. Hospitalisation with a primary diagnosis of heart failure (HHF)
3. All-cause mortality

### Acquisition and linkage of outcome data

Clinical events in New Zealand public and private hospitals are entered into the National Minimum Dataset (NMDS), whilst information concerning mortality is recorded in the Mortality Collection. Both code events according to the International Statistical Classification of Diseases and Related Health Problems (10^th^ Revision, Australian Modification; ICD-10- AM) and can be linked to an individual patient using a unique NHI number. Information concerning discharges and interventions at public and private hospitals between January 1^st^ 2010 and August 31^st^ 2022 (including NHI number, date of admission/procedure, primary diagnosis or procedure code, ethnicity, date of birth, and sex), plus mortality (date and primary cause of death) over the same time period were provided by the Ministry of Health and linked to each participant’s echocardiographic data using their unique NHI number.

### Study approval

Consultation with Māori was undertaken with the Ngāi Tahu Research Consultation Committee. The study received ethical approval from the New Zealand Central Health and

Disability Ethics Committee (ref: 21/CEN/15) and locality approval from Te Whatu Ora – Southern.

### Statistical analysis

All analyses were performed on a de-identified dataset with NHI numbers replaced by anonymous identifiers. Continuous data are expressed as mean (standard deviation) if normally distributed, and otherwise as median (interquartile range). Data were analysed using the Mann-Whitney *U*-test if continuous and non-normally distributed, and with ANOVA if normally distributed. Categorical variables were analysed using the Chi-square test.

Kaplan-Meier survival curves were produced for the primary and secondary outcomes, and stratified according to severity of CAVD. Group differences were assessed using the log-rank test with Benjamini-Hochberg correction for multiple testing. Assessment of the proposed causal diagrams demonstrated that estimation of the effect of CAVD on outcomes was not possible (due to lack of information concerning cardiovascular risk factors). This particular analysis was therefore restricted to descriptive statistics. All analyses (including data cleaning) were performed using RStudio with R version 3.6.3 (9, 10).

## Results

The final cohort numbered 24,699 people after exclusion of those with missing CAVD status and an LV mass >1000g. Of these, 8,066 had aortic sclerosis, 920 had mild AS, and 531 had moderate AS. Patients were followed up for a median of 8.1 years (IQR: 5.6 – 10.5, maximum 12.6 years) and 7,898 (38.8%) of those with at least five years follow-up (n=20,371) experienced the primary composite outcome (all-cause mortality, n=7,246; HHF, n=652), and 2,106 (35.6%) experienced HHF. Amongst those with moderate AS and at least five years follow-up (n=436), 267 (61.2%) experienced the primary outcome (all-cause mortality, n=254; heart failure hospitalisation, n=84, with all but 13 of these subsequently dying). Similarly, in mild AS (n=720), 416 (57.8%) experienced the primary outcome (all-cause mortality, n=383; heart failure hospitalisation, n=118, with all but 33 of these subsequently dying).

Worsening stage of cardiac damage was associated with an increased rate of the primary outcome for both mild and moderate AS (**Figure 1** and **Figure 2**, **Table 2** and **Table 3**).

**Figure 1:**
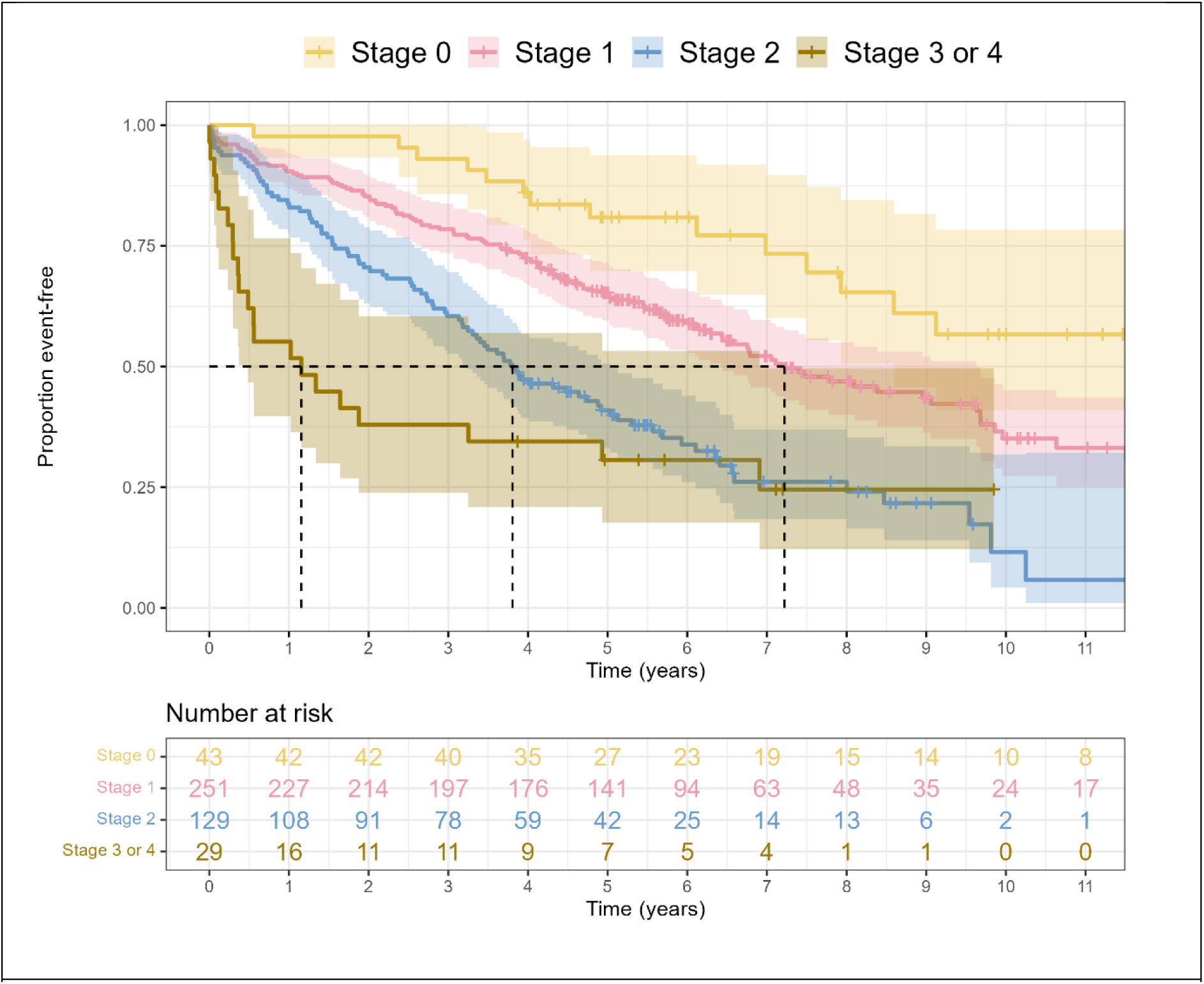
Kaplan-Meier curve of mortality or heart failure hospitalisation in moderate AS, stratified according to stage of cardiac damage.

**Figure 2:**
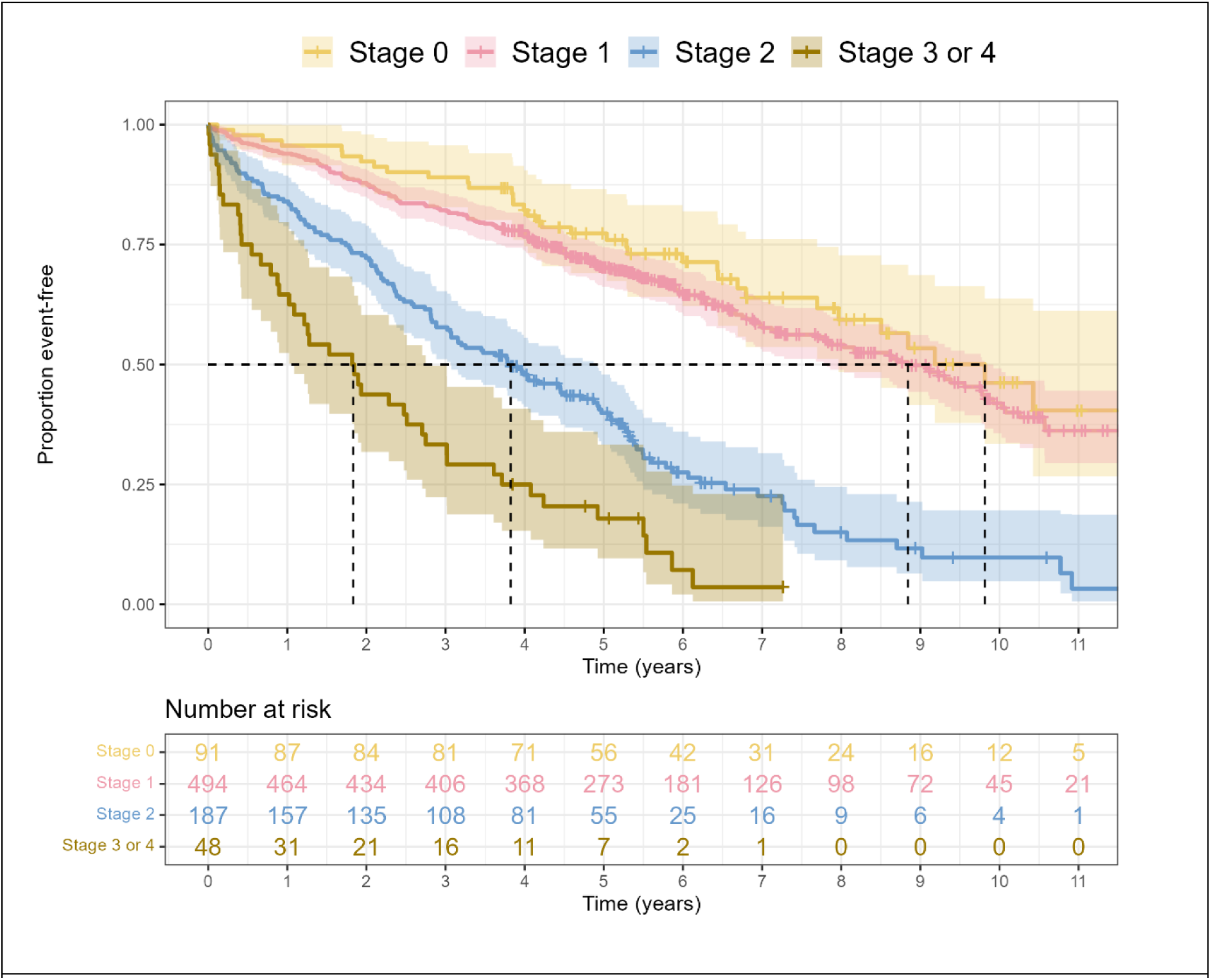
Kaplan-Meier curve of mortality or heart failure hospitalisation in mild AS, stratified according to stage of cardiac damage.

**Table 1:** Baseline characteristics of the cohort for survival analysis. Cells are formatted as n (%) unless otherwise specified. Abbreviations: AVI, aortic valve intervention; SD, standard deviation; MELAA, Middle Eastern, Latin American, or African; CAVD, calcific aortic valve disease

|  | AVI<br>(N=552) | No CAVD<br>(N=14257) | Stage 0<br>(N=1379) | Stage 1<br>(N=5157) | Stage 2<br>(N=2583) | Stage 3<br>(N=291) | Stage 4<br>(N=480) | Overall<br>(N=24699) |
| --- | --- | --- | --- | --- | --- | --- | --- | --- |
| **Age (years)** |  |  |  |  |  |  |  |  |
| Mean (SD) | 69.90<br>(14.10) | 56.80<br>(16.80) | 70.90<br>(10.80) | 72.70<br>(11.30) | 77.00<br>(9.51) | 81.20<br>(8.53) | 75.00<br>(11.60) | 63.90<br>(17.00) |
| **Sex** |  |  |  |  |  |  |  |  |
| Female | 184<br>(33.3) | 6875<br>(48.2) | 636 (46.1) | 2490<br>(48.3) | 1012<br>(39.2) | 180<br>(61.9) | 163 (34.0) | 11540<br>(46.7) |
| Male | 368<br>(66.7) | 7382<br>(51.8) | 743 (53.9) | 2667<br>(51.7) | 1571<br>(60.8) | 111<br>(38.1) | 317 (66.0) | 13159<br>(53.3) |
| **Ethnicity** |  |  |  |  |  |  |  |  |
| European | 524<br>(94.9) | 12549<br>(88.0) | 1294<br>(93.8) | 4836<br>(93.8) | 2434<br>(94.2) | 263<br>(90.4) | 423 (88.1) | 22323<br>(90.4) |
| Māori | 21 (3.8) | 915 (6.4) | 53 (3.8) | 178 (3.5) | 95 (3.7) | 12 (4.1) | 38 (7.9) | 1312 (5.3) |
| Pacific Peoples | 6 (1.1) | 255 (1.8) | 7 (0.5) | 35 (0.7) | 19 (0.7) | 8 (2.7) | 11 (2.3) | 341 (1.4) |
| Asian | 0 (0.0) | 350 (2.5) | 12 (0.9) | 52 (1.0) | 12 (0.5) | 5 (1.7) | 3 (0.6) | 434 (1.8) |
| MELAA | 0 (0.0) | 96 (0.7) | 6 (0.4) | 17 (0.3) | 2 (0.1) | 0 (0.0) | 0 (0.0) | 121 (0.5) |
| Other | 1 (0.2) | 92 (0.6) | 7 (0.5) | 39 (0.8) | 21 (0.8) | 3 (1.0) | 5 (1.0) | 168 (0.7) |
| **Aortic valve maximum velocity (m/s)** |  |  |  |  |  |  |  |  |
| Mean (SD) | 2.07<br>(1.50) | 1.23 (0.54) | 1.66 (0.79) | 1.76 (1.04) | 1.78 (1.18) | 1.84<br>(1.12) | 1.45 (0.90) | 1.46 (0.86) |
| Not reported | 16 (3.0) | 1834<br>(14.8) | 113 (8.9) | 282 (5.8) | 177 (7.4) | 20 (7.4) | 58 (13.7) | 2500 (11.3) |
| **CAVD severity** |  |  |  |  |  |  |  |  |
| No CAVD | 0 (0.0) | 14257<br>(100.0) | 0 (0.0) | 0 (0.0) | 0 (0.0) | 0 (0.0) | 0 (0.0) | 14257<br>(57.7) |
| Sclerosis | 0 (0.0) | 0 (0.0) | 1208<br>(87.6) | 4187<br>(81.2) | 2021<br>(78.2) | 228<br>(78.4) | 422 (87.9) | 8066 (32.7) |
| Mild | 0 (0.0) | 0 (0.0) | 106 (7.7) | 532 (10.3) | 230 (8.9) | 24 (8.2) | 28 (5.8) | 920 (3.7) |
| Moderate | 0 (0.0) | 0 (0.0) | 50 (3.6) | 285 (5.5) | 165 (6.4) | 17 (5.8) | 14 (2.9) | 531 (2.1) |
| Severe | 0 (0.0) | 0 (0.0) | 15 (1.1) | 153 (3.0) | 167 (6.5) | 22 (7.6) | 16 (3.3) | 373 (1.5) |
| AVR | 552<br>(100.0) | 0 (0.0) | 0 (0.0) | 0 (0.0) | 0 (0.0) | 0 (0.0) | 0 (0.0) | 552 (2.2) |
| **Mitral annular calcification** |  |  |  |  |  |  |  |  |
| Yes | 185<br>(33.5) | 627 (4.4) | 250 (18.1) | 1266<br>(24.5) | 836 (32.4) | 109<br>(37.5) | 102 (21.2) | 3375 (13.7) |
| No | 361<br>(65.4) | 13386<br>(93.9) | 1109<br>(80.4) | 3836<br>(74.4) | 1715<br>(66.4) | 179<br>(61.5) | 372 (77.5) | 20958<br>(84.9) |
| Not reported | 6 (1.1) | 244 (1.7) | 20 (1.5) | 55 (1.1) | 32 (1.2) | 3 (1.0) | 6 (1.2) | 366 (1.5) |
| **Tricuspid regurgitation** |  |  |  |  |  |  |  |  |
| None | 297<br>(53.8) | 10098<br>(70.8) | 1083<br>(78.5) | 3839<br>(74.4) | 1425<br>(55.2) | 12 (4.1) | 109 (22.7) | 16863<br>(68.3) |
| Mild | 120<br>(21.7) | 1084 (7.6) | 164 (11.9) | 598 (11.6) | 923 (35.7) | 34<br>(11.7) | 194 (40.4) | 3117 (12.6) |
| Moderate | 27 (4.9) | 150 (1.1) | 0 (0.0) | 0 (0.0) | 0 (0.0) | 217<br>(74.6) | 119 (24.8) | 513 (2.1) |
| Severe | 3 (0.5) | 12 (0.1) | 0 (0.0) | 0 (0.0) | 0 (0.0) | 23 (7.9) | 21 (4.4) | 59 (0.2) |
| Not reported | 105<br>(19.0) | 2913<br>(20.4) | 132 (9.6) | 720 (14.0) | 235 (9.1) | 5 (1.7) | 37 (7.7) | 4147 (16.8) |
| **Bicuspid aortic valve** |  |  |  |  |  |  |  |  |
| Yes | 0 (0.0) | 100 (0.7) | 31 (2.2) | 139 (2.7) | 41 (1.6) | 1 (0.3) | 8 (1.7) | 320 (1.3) |
| No | 552<br>(100.0) | 14157<br>(99.3) | 1348<br>(97.8) | 5018<br>(97.3) | 2542<br>(98.4) | 290<br>(99.7) | 472 (98.3) | 24379<br>(98.7) |
| **Cardiac rhythm** |  |  |  |  |  |  |  |  |
| Sinus rhythm | 306<br>(55.4) | 10805<br>(75.8) | 1268<br>(92.0) | 4314<br>(83.7) | 1124<br>(43.5) | 103<br>(35.4) | 212 (44.2) | 18132<br>(73.4) |
| Atrial fibrillation or flutter | 77<br>(13.9) | 1073 (7.5) | 1 (0.1) | 29 (0.6) | 1176<br>(45.5) | 145<br>(49.8) | 206 (42.9) | 2707 (11.0) |
| Heart block | 4 (0.7) | 99 (0.7) | 8 (0.6) | 61 (1.2) | 21 (0.8) | 1 (0.3) | 5 (1.0) | 199 (0.8) |
| Other | 44 (8.0) | 202 (1.4) | 7 (0.5) | 122 (2.4) | 83 (3.2) | 10 (3.4) | 22 (4.6) | 490 (2.0) |
| Not reported | 121<br>(21.9) | 2078<br>(14.6) | 95 (6.9) | 631 (12.2) | 179 (6.9) | 32<br>(11.0) | 35 (7.3) | 3171 (12.8) |

| LV systolic impairment |  |  |  |  |  |  |  |  |
| --- | --- | --- | --- | --- | --- | --- | --- | --- |
| Normal | 373<br>(67.6) | 11215<br>(78.7) | 1222<br>(88.6) | 3794<br>(73.6) | 1475<br>(57.1) | 164<br>(56.4) | 136 (28.3) | 18379<br>(74.4) |
| Hyperdynamic | 7 (1.3) | 159 (1.1) | 35 (2.5) | 78 (1.5) | 28 (1.1) | 1 (0.3) | 7 (1.5) | 315 (1.3) |
| Mild | 78<br>(14.1) | 1163 (8.2) | 27 (2.0) | 539 (10.5) | 364 (14.1) | 33<br>(11.3) | 49 (10.2) | 2253 (9.1) |
| Moderate | 44 (8.0) | 642 (4.5) | 3 (0.2) | 257 (5.0) | 279 (10.8) | 30<br>(10.3) | 63 (13.1) | 1318 (5.3) |
| Severe | 22 (4.0) | 422 (3.0) | 1 (0.1) | 106 (2.1) | 179 (6.9) | 22 (7.6) | 130 (27.1) | 882 (3.6) |
| Not reported | 28 (5.1) | 656 (4.6) | 91 (6.6) | 383 (7.4) | 258 (10.0) | 41<br>(14.1) | 95 (19.8) | 1552 (6.3) |

**Table 2:** Cox proportional hazards model for mortality or heart failure hospitalisation in moderate AS according to stage of cardiac damage. Reference value: Stage 0.

|  | Univariate |  | Age and sex adjusted |  |
| --- | --- | --- | --- | --- |
|  | Hazard ratio | P value | Hazard ratio | P value |
| Stage 1 | 2.00 (1.15 – 3.48) | <0.05 | 1.61 (0.92 – 2.81) | 0.09 |
| Stage 2 | 4.03 (2.28 – 7.12) | <0.001 | 2.71 (1.53 – 4.81) | <0.001 |
| Stage 3 or 4 | 5.81 (2.94 – 11.48) | <0.001 | 3.97 (1.99 – 7.95) | <0.001 |

**Table 3:**
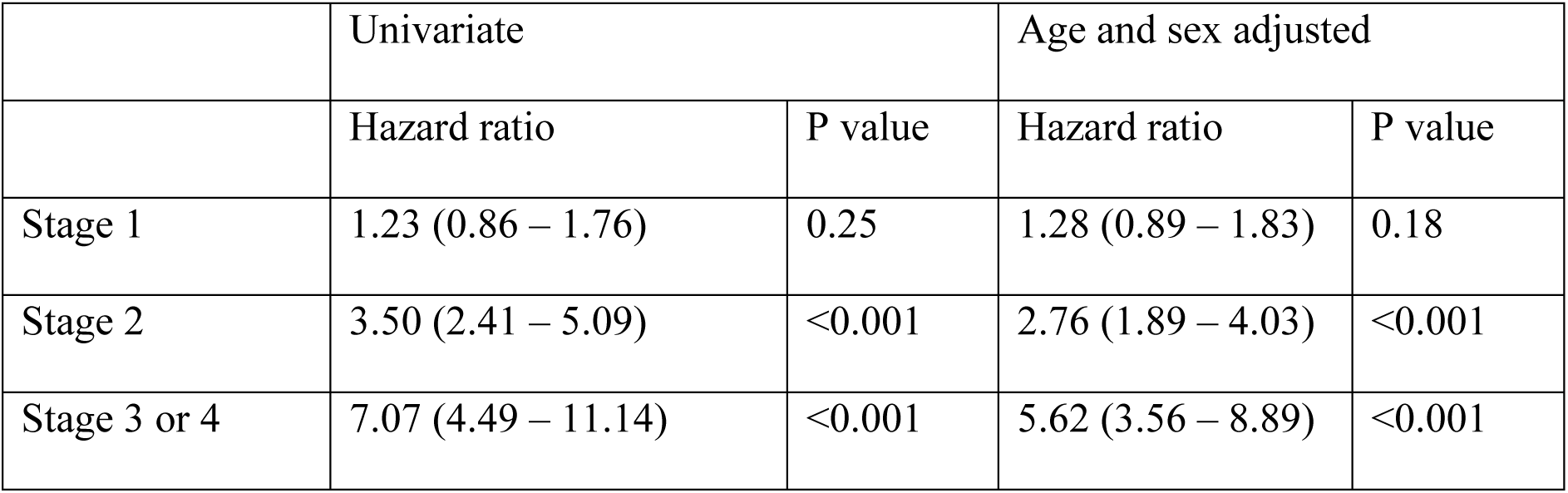
Cox proportional hazards model for mortality or heart failure hospitalisation in mild AS according to stage of cardiac damage. Reference value: Stage 0.

### Mortality censored at the time of aortic valve intervention (AVI)

To remove the potential effect of AVI on mortality, Kaplan-Meier curves of mortality stratified according to stage of cardiac damage and censored at time of AVI or completion of follow-up are presented for both moderate (**Figure 3A**) and mild AS (**Figure 3B)**. The risk of mortality increased according to the stage of cardiac damage, even before any AVI. Similarly, Cox proportional hazards models are reported in **Table 4** for mortality stratified according to stage of cardiac damage and censored at the time of AVI or completion of follow up. All stages of cardiac damage were significantly associated with mortality in moderate AS (compared to Stage 0), whereas stages 2, 3 and 4 cardiac damage were significantly associated with mortality in mild AS.

**Figure 3:**
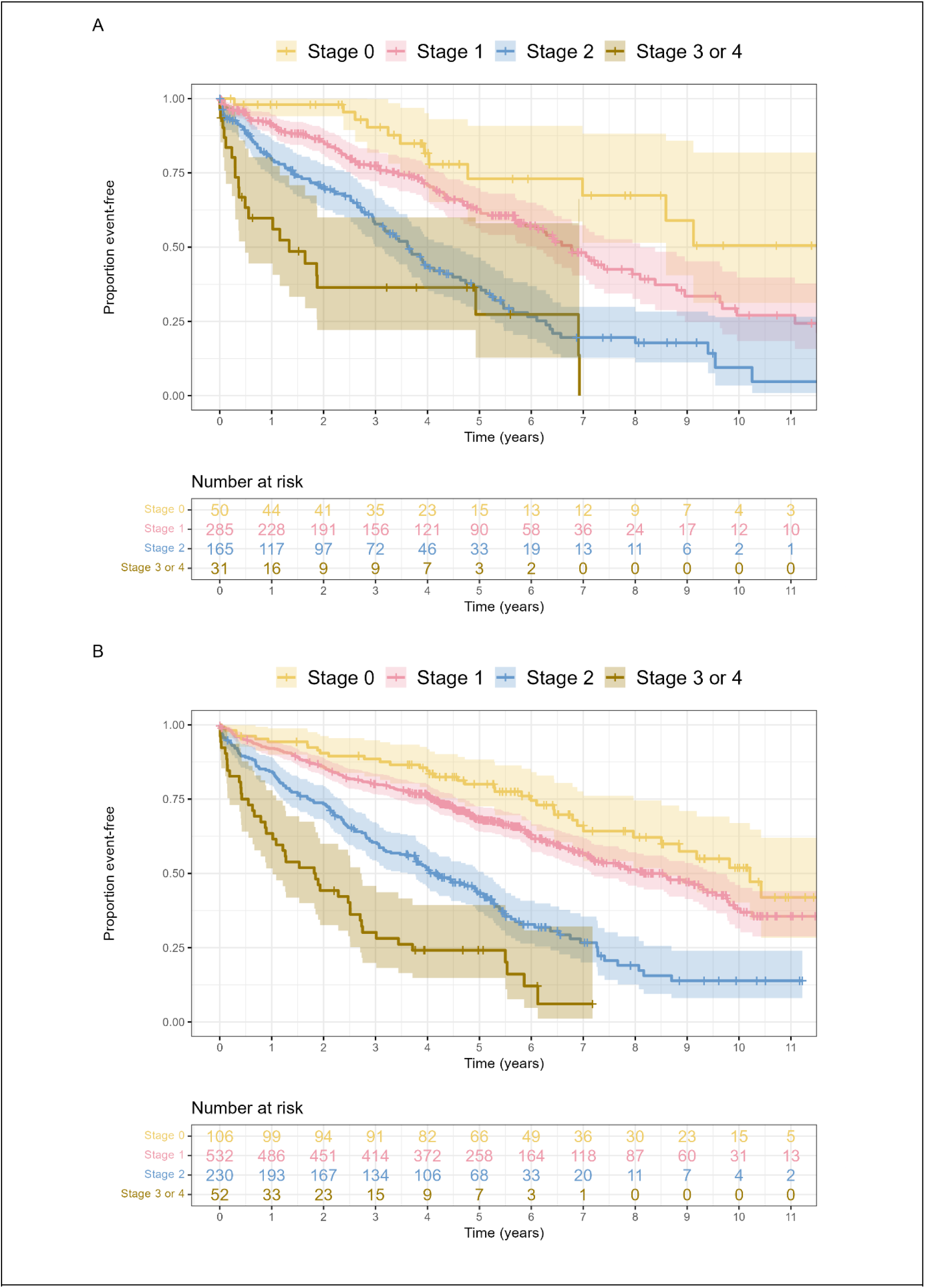
Kaplan-Meier curves of mortality stratified according to stage of cardiac damage in (A) moderate AS, and (B) mild AS, censored at the time of AVI or completion of follow-up.

**Table 4:** Cox proportional hazards model for mortality in mild or moderate AS stratified according to stage of cardiac damage, adjusted for age and sex, and censored at the time of AVI or completion of follow-up. Reference level: Stage 0.

|  | **Hazard ratio** | **95% confidence interval** | **P value** |
| --- | --- | --- | --- |
| **Moderate AS** |  |  |  |
| Stage 1 | 1.99 | 1.09 – 3.61 | 0.024 |
| Stage 2 | 4.18 | 2.29 – 7.63 | <0.001 |
| Stage 3/4 | 7.01 | 3.43 – 14.33 | <0.001 |
| **Mild AS** |  |  |  |
| Stage 1 | 1.40 | 0.98 – 1.99 | 0.06 |
| Stage 2 | 3.22 | 2.22 – 4.66 | <0.001 |
| Stage 3/4 | 6.87 | 4.35 – 10.84 | <0.001 |

## Discussion

In this large study of over 20,000 patients undergoing clinically indicated echocardiography, we examined a primary outcome of frequency of hospitalisation with heart failure or mortality in those with moderate and mild AS. Cardiac damage was common in both cohorts and increasing levels were significantly associated with the primary outcome after adjustment for age and sex. Furthermore, similiar observations were seen even prior to any AVI.

Staging systems to quantify and assess the impact of cardiac damage associated with CAVD were first conceived in 2017(1) and previous studies have generally focused on their utility in cohorts with severe AS. In comparison with another study restricted to moderate AS patients, our cohort had a slightly lower prevalence of stage 3 or 4 cardiac damage (3.2% and 2.6% compared to 10.6% and 6.9%, respectively) (11), but a greater hazard ratio for mortality across all stages of cardiac damage (for instance, 7.01 compared with 4.46 for Group 4 vs.

Group 0) (11). Unfortunately, our dataset did not contain information concerning other cardiovascular risk factors, thereby making further comparisons challenging. Risk factors and comorbidities almost certainly confound the relationship between the stage of cardiac damage and all-cause mortality. Our data demonstrate that each incremental stage of cardiac damage was associated with a HR of 1.62 for the composite outcome, exceeding the ratios of 1.31 and 1.45 reported previously in patients with moderate to severe AS (1, 2).

A key question of any staging system for cardiac damage associated with CAVD is whether it can better identify patients who will benefit from AVI (and when such intervention should take place). Evidence from clinical cohorts in Australia and the United States suggest that moderate AS is a subgroup requiring closer surveillance given higher observed mortality rates compared to mild or no AS (12, 13). Current guidelines do not recommend AVI in moderate AS, despite observational data suggesting that early AVI is associated with a 45% reduction in the risk of all-cause mortality this group (14, 15). However, such data are potentially confounded by the fact that those who undergo AVI are far more likely to have fewer medical comorbidities that impact upon survival. Randomised controlled trials have demonstrated the benefit of early surgery in patients with asymptomatic severe AS and normal left ventricular function (16) and ongoing trials, such as the PROGRESS trial of AVI vs medical surveillance, are addressing the same question in patients with moderate AS (17).

### Mild AS

To our knowledge, our study is the first to demonstrate that mortality is linked to the stage of cardiac damage in mild AS, and it is noteworthy that the magnitude of this association was not significantly different between mild and moderate AS. The haemodynamic impact of mild AS is unlikely to be sufficient to produce Stage 3 or Stage 4 cardiac damage, and other comorbidities are therefore likely to be influential.

### Utility of the staging system

Our data suggest that the stage of cardiac damage predicts prognosis in both moderate and mild AS to a similar extent. If the haemodynamic consequences of AS were solely responsible for this association, then it would stand to reason that worsening haemodynamic profiles would result in higher event rates. However, our observations do not support this argument, suggesting that outcomes may not be directly attributable to the haemodynamic consequences of valve disease and are unlikely to be restored to by AVI alone. Nevertheless, the prognostic information provided by systems for the staging of cardiac damage may be useful to identify patients at particular risk, direct the introduction of optimal medical therapy and determine the mode and timing of AVI.

## Strengths and limitations

To our knowledge, this is the first study to apply the concept of cardiac damage staging to a large number of patients with mild and moderate AS, and to examine non-mortality outcomes such as heart failure hospitalisation. A particular strength of our study is the large number of patients involved with prolonged follow up (median 8 years) and a sufficient number of clinical events to provide robust analysis. Furthermore, the likelihood of “missed” events is low, given New Zealand’s centralised hospital reporting system and the inclusion of private hospital data. Our study also drew on a complete echocardiographic dataset and is therefore representative of the population undergoing clinically indicated investigations. However, we lacked information on cardiovascular risk factors, which potentially confound the relationship between valve disease and prognostic outcome, and would help to further elucidate the connection between cardiac damage and adverse events. Information on the progression of AS was similarly not available and thus could not be accounted for. Finally, the numbers of patients with stage 3 or 4 cardiac damage were relatively small and these groups were therefore combined for analysis, thereby limiting our ability to examine the difference in event rates between them.

## Conclusions

The stage of cardiac damage associated with CAVD predicts prognosis to a similar extent in both moderate and mild AS, suggesting that this association is at least partly independent of haemodynamic effects and may not be completely reversible by AVI alone. Nevertheless, systems for the staging of cardiac damage provide a clear delineation of risk that may facilitate clinical decision making concerning the implementation of medical therapy and the timing and mode of AVI. Aortic stenosis is a disease of the valve and myocardium, and enhanced strategies to identify and treat cardiac damage in conjunction with AVI are now required.

## Authors’ contribution

MKM developed the protocol, performed the measurements and analysis, and wrote the first draft of the manuscript. SC and GJT conceived the study and contributed to the protocol design. All authors contributed to the analysis of results, and reviewed and approved the final manuscript.

## Data Availability

The data underlying this article cannot be shared publicly on account of the ethics approval conditions of this study.

